# Improved Sleep and Clinical Outcomes Following a Single-Session Intervention among Adolescents with Borderline Personality Disorder Features

**DOI:** 10.64898/2026.09.22.26363695

**Authors:** Erin A. Kaufman, Sarah E. Victor, Jin H. Prunuske, Jenny L. Nguyen, Geoffrey Laforge

**Affiliations:** Department of Psychiatry & Huntsman Mental Health Institute, Spencer Fox Eccles School of Medicine, University of Utah, 383 Colorow Dr., Salt Lake City, UT, 84108 USA; Department of Psychological Sciences, Texas Tech University, 2700 18th St., Lubbock, TX 79410 USA; Department of Neurology, University of Utah, Salt Lake City, UT, 383 Colorow Dr., Salt Lake City, UT, 84108 USA

**Keywords:** Borderline personality disorder, adolescents, intervention, sleep, ecological momentary assessment, EEG

## Abstract

Borderline personality disorder (BPD) is a serious mental illness that often emerges in adolescence. Although efficacious treatments have been developed, they are often lengthy, expensive, inaccessible, and require specialized training to deliver. Optimizing BPD prevention and early treatment is an important public health priority. Disrupted sleep is closely linked to key BPD features, and prior research has documented a range of sleep problems among BPD samples that appear to worsen symptom course and duration. This intervention trial (NCT04507308) assessed the effects of a single-session sleep-focused treatment on adolescent sleep and BPD-related symptoms. Fifty-seven adolescents (ages 13-18) with 3+ clinically impairing BPD features and at least some self-reported sleep-related difficulties participated with a caregiver. Participants completed baseline surveys, interviews, and 10-day ecological momentary assessment (EMA) protocol to examine BPD symptoms and sleep in daily life. Youth also collected at-home ambulatory electroencephalogram (EEG) recordings before undergoing the treatment informed by the Transdiagnostic Sleep and Circadian Intervention. Post-treatment, youth practiced their new sleep routine for approximately one month before undergoing reassessment (EMA, EEG, surveys). Many indices of sleep quantity and quality improved across both objective (EEG) and subjective assessments (EMA and self- and caregiver-report). Youth reported an earlier clock time at sleep onset and earlier waketime, more sleep, and fewer awakenings in daily life. Every sleep-related questionnaire score improved at post-intervention and was maintained at one-month follow-up. Multiple (though not all) indices of BPD symptoms also improved significantly. Findings suggest sleep may be a promising intervention target for at-risk youth.

---

Borderline personality disorder (BPD) is a persistent and profoundly impairing mental health condition characterized by sustained dysregulation across multiple core domains of functioning (e.g., interpersonal turmoil, affective lability, identity disturbance, and impulsivity; American Psychiatric Association [APA], 2022). BPD is associated with severe and often chronic impairments in psychosocial functioning (Gunderson, 2011; Leichsenring et al., 2023; Zanarini et al., 2010), high service utilization, and elevated financial costs (Bode et al., 2017; Hastrup et al., 2022; Wagner et al., 2022). Approximately 69% to 80% of persons with this condition engage in self-injurious behavior with as many as 6-9% dying by suicide (Lak et al., 2025; Temes et al., 2019; Zanarini et al., 2005). BPD often first manifests in adolescence and even subthreshold symptoms are associated with significant functional impairment and morbidity (Brager-Larsen et al., 2023; Zimmerman et al., 2013). Importantly, although efficacious treatment approaches exist (e.g., Dialectical Behavior Therapy [DBT]; Linehan, 2025; Rathus & Miller, 2015), available interventions are typically expensive, lengthy, intensive, and difficult to access (Tusiani-Eng & Yeomans, 2018). Furthermore, up to 50% of persons with this condition do not respond to frontline therapy (Woodbridge et al., 2022). Identifying methods to optimize BPD prevention and treatment is an urgent public health priority.

Disrupted sleep is closely linked to defining characteristics of BPD such as higher sensitivity and reactivity to stress, behavioral impulsivity, interpersonal dysfunction, and poor emotion regulation (Beattie et al., 2015; Ben Simon & Walker, 2018; Demos et al., 2016; Saksvik-Lehouillier et al., 2020; Winsper et al., 2017; Yoo et al., 2023). Studies have documented a range of sleep disturbances in adult BPD samples and youth with BPD features, and have demonstrated that sleep problems interact with, but are not better accounted for, by concomitant depression or other psychiatric disorders (Jenkins et al., 2022; King et al., 2025; Mendoza Alvarez et al., 2025; Remeeus et al., 2024; Selby et al., 2013; Winsper et al., 2017). Importantly, sleep problems may predispose persons to BPD (Durdurak et al., 2022), worsen BPD symptom course and chronicity (Plante et al., 2013), heighten risk of suicide (Balestrieri et al., 2006; DeShong, et al., 2019; Kaurin, et al., 2022), and interfere with cognitive processes that are vital to behavioral intervention (e.g., memory consolidation and attentional processes; Cellini et al., 2017; Curcio, et al., 2006; García et al., 2021; Gobin et al., 2015). A recent daily-diary study with adults found that participants with higher BPD features exhibited greater negative affect on days when they reported worse sleep quality, as compared to participants with lower BPD features (Klein et al., 2026). A greater understanding of sleep disturbance in BPD may help enrich treatment protocols, which currently place limited emphasis on sleep difficulties. Although approaches like Cognitive Behavior Therapy for insomnia (CBT-I; Walker et al., 2022) and the youth version of the Transdiagnostic Sleep and Circadian Intervention (TSC-Youth; Dolsen et al., 2023; Harvey, 2016; Harvey & Buysse, 2017) are effective with many populations, it is currently unknown whether sleep-focused interventions can be tolerated or followed adherently by youth with significant BPD features.

The present clinical trial sought to assess the preliminary utility of a single-session sleep intervention for improving sleep quality and BPD symptoms among adolescents with BPD features. Randomized controlled trials of single-session interventions for youth and adult mental health problems have previously yielded small to medium effect sizes and significant clinical utility relative to control conditions (see Schleider et al., 2025 and Schleider & Weisz, 2017 for reviews and meta-analyses) and some preliminary evidence has emerged for a single-session sleep-focused intervention for adolescents with irregular or insufficient sleep (see Levenson et al., 2025; Wescott et al., 2026). For this study, we examined potential changes in self-, informant-reported, and objective indices of sleep, as well as self-reported BPD symptoms as a function of sleep training (pre-post intervention effects). We hypothesized adolescent sleep would improve across measures of self- and caregiver-reported sleep disturbance and sleep-related impairment, and electroencephalogram (EEG) derived measures of nightly sleep at home following the intervention. We also expected reductions in adolescent BPD symptoms, and improved mood stabilization post-intervention.

## Method

### Participants

The 57 caregiver-adolescent dyads in the current study were recruited from June 2024 through March 2026 (NCT04507308; distribution by month reported in supplemental material). Based on prior single-session intervention studies (Schleider et al., 2025; Schleider & Weisz, 2017), we anticipated effect sizes in the medium range, which prior work suggested was feasible to obtain in a sample of this size (Levenson et al., 2025; Wescott et al., 2026). The vast majority of participants were recruited following outreach by the study team (see Figure 1). Research assistants performed electronic health record review of the University of Utah’s emergency department, and the Huntsman Mental Health Institute’s adolescent inpatient unit and their intensive outpatient programming. Caregivers with an appropriately aged youth presenting to treatment for self-inflicted injury, suicide attempt, disruptive mood dysregulation disorder, behavioral problems or aggression, BPD, or borderline personality features were contacted and invited to screen for the study. Most families had signed a consent form upon admission/enrollment that indicated interest in being contacted for research purposes^1^. Additional families expressed interest following advertisements placed in a Salt Lake City community newsletter and the University of Utah’s Study Locator Webpage. Adolescents and their caregivers (either a parent or a grandparent who was a legal guardian) were screened by phone or Zoom for eligibility. Participant flow and additional details regarding recruitment source is illustrated in Figure 1.

**Figure 1:**
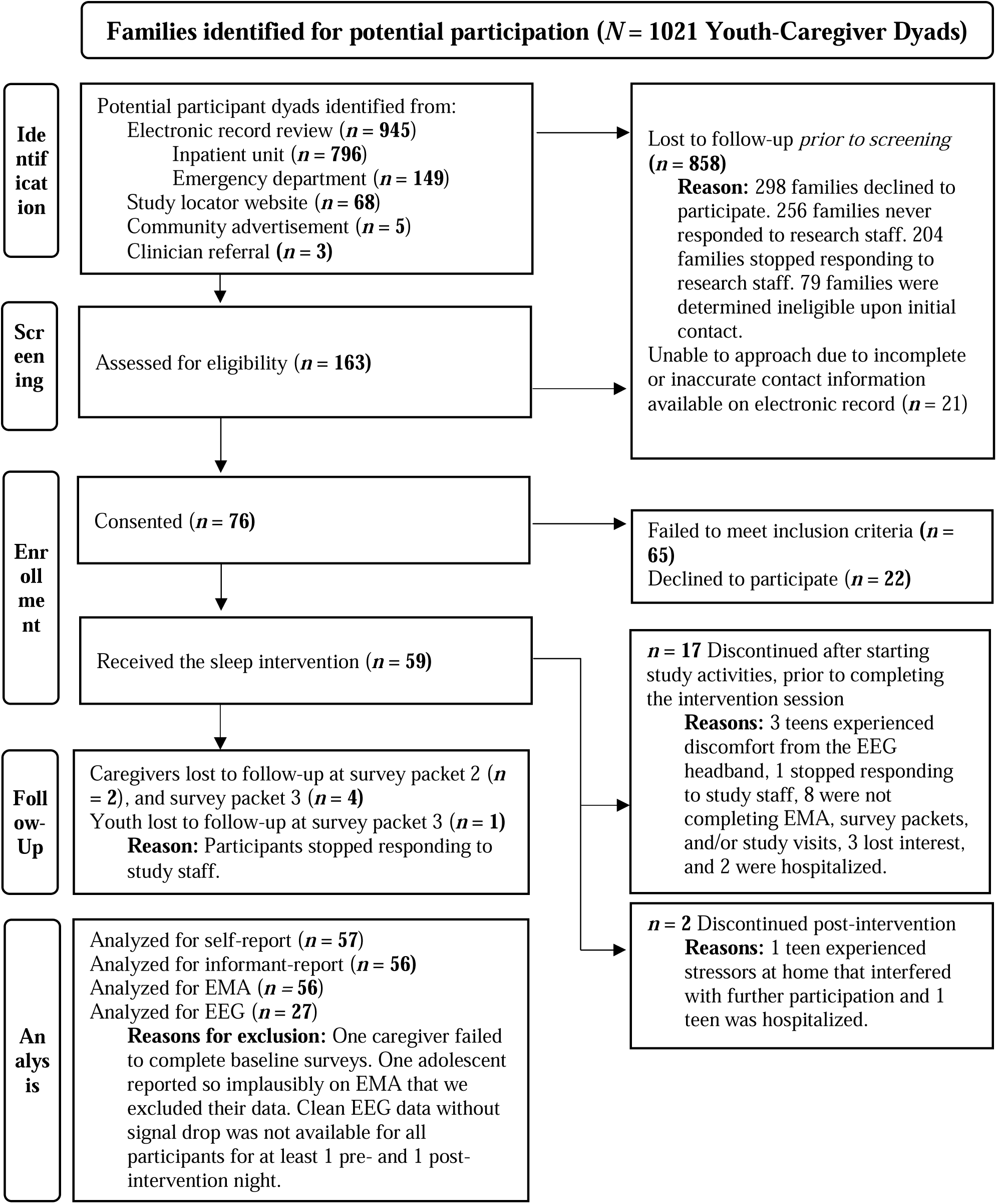
CONSORT diagram illustrating the flow of participants through the study.

Adolescents were: (a) between the ages of 13 and 18 at the time of enrollment, (b) experiencing 3+ clinically significant BPD symptoms, (c) experiencing at least some self-reported sleep difficulties and/or impairment, (d) fluent in English, (e) and able to consistently access to a smart-phone or email to complete ecological momentary assessment (EMA) surveys. Exclusion criteria were youth intellectual disability, pervasive developmental disorder, or schizophrenia spectrum diagnosis, and/or not having a legal guardian available for participation with English language proficiency. All study activities were approved by the University of Utah Institutional Review Board. Caregivers provided informed consent and permission for youth under the age of 18 to participate. Adolescents provided informed assent/consent depending on their age. Youth were compensated up to $165 for participation. To enhance EMA adherence, participants were compensated based on the percentage of total surveys completed. Adolescents who completed 85% or more of their assessments received a $20 bonus payment.

### Measures

#### Screening instruments

Research assistants read potential participants a script that briefly described study tasks and the topics queried in the screening interview. Study personnel received verbal permission to proceed (guardian permission was obtained prior to speaking with adolescents below the age of 18). Caregivers and adolescents were each separately asked about demographics, their willingness and ability to participate, adolescents’ current medical conditions, medications, and history of exclusionary mental health conditions.

To assess eligibility, youth also completed a semi-structured interview version of the 10-item *McLean Screening Instrument for Borderline Personality Disorder* (MSI-BPD; Zanarini et al., 2003), adapted such that interviewers asked follow-up questions to determine frequency, severity, chronicity, pervasiveness, and functional impairment associated with each BPD criteria. Youth also completed short-forms of the *Patient-Reported Outcomes Measurement Information System Pediatric Sleep Disturbances* (PROMIS-PSD) and *Pediatric Sleep-related Impairment* item banks (PROMIS-PSRI; Forrest et al., 2018) to ensure participants were experiencing at least some level of sleep related difficulty in the past week that could be targeted with intervention. Internal consistency reliabilities in the current sample and additional details regarding scoring for all administered questionnaires are available in the supplemental material.

#### Survey Packets

Eligible participants completed online survey packets at baseline (pre-treatment), following 3-4 weeks of intervention practice (post-treatment), and at a 1-month follow-up.

##### Caregiver Surveys

At baseline, caregivers completed demographic questions about themselves and their child drawn from the National Center for Advancing Translational Sciences (n.d.), as well as the *Child Behavior Checklist* to characterize youth psychopathology (CBCL; Achenbach, 2009; Achenbach & Rescorla, 2001). The CBCL is a 113-item measure and one of the most used and widely validated measures of youth behavior problems (Achenbach, 2009). Prior research has yielded acceptable construct validity for internalizing (*r* = .56 to .72) and externalizing behaviors (*r* = .52 to .88; Achenbach, 1991), and acceptable to high internal consistency (_α_ = .68 –.92 for all behavioral scales in Achenbach, 1991; Cronbach’s _α_ = .93–.95 for internalizing and externalizing scales; Kaufman et al., 2016). To assess their children’s sleep-related functioning, caregivers completed companion parent versions of the PROMIS-PSD and a 4-item version of the PROMIS-PSRI (Forrest et al., 2018) at all three survey time points.

##### Adolescent Surveys

At baseline, youth completed demographic questions and self-report scales chosen to assess relevant psychopathology and sleep. Adolescents completed the *Level 2 PROMIS Emotional Distress—Depression—Pediatric* Item Bank (PROMIS-EDDP; APA, 2013) given the common co-occurrence and symptom overlap between BPD and depression (Kew et al., 2025). This 14-item Likert-style questionnaire assesses depression severity in the prior week, with raw sums converted to t-scores for interpretation (Irwin et al., 2010). The PROMIS-EDDP was included to help characterize youth psychopathology.

Multiple measures were used to assess BPD features. We administered the *Borderline Personality Features Scale for Children-11* (BPFSC-11; Sharp et al., 2014) at baseline and post-intervention time points. This brief self-report scale has previously demonstrated excellent internal consistency (Cronbach’s alpha = 0.87) and high convergent validity (Aune et al., 2025). BPFSC-11 items primary assess perceived interpersonal disconnection and negative relationships (i.e., feeling lonely; wanting to let others know how much they’ve hurt the respondent; being let down by close others; being “really mean” with friends; worrying others will leave and not come back). Two items focus on emotion dysregulation (i.e., experiencing really strong emotions; going back and forth between different feelings), two on difficulties with identity (i.e., feeling something important is missing about the respondent, but not knowing what it is; experiencing frequent changes in how the respondent feels about themself) and two on impulsivity or carelessness (i.e., being careless with important things; getting into trouble for doing things without thinking). In order to examine intervention effects in greater depth, we also administered validated instruments tapping core features of BPD. These include: the 18-item *Difficulties in Emotion Regulation Scale Short Form* as a measure of emotion dysregulation (DERS-SF; Kaufman et al., 2016), the 20-item short version of the *Urgency, Premeditation (lack of), Perseverance (lack of), Sensation Seeking, Positive Urgency, Impulsive Behavior Scale* as a measure of impulsivity (UPPS-P; Cyders et al., 2007; Whiteside & Lynam, 2001), the 27-item *Self-Concept and Identity Measure* as a measure of identity disturbance (SCIM; Kaufman et al., 2015), and the 15-item *Suicide Ideation Questionnaire Junior* (SIQ-Jr.; Reynolds, 1988) as a measure of suicide risk (Courtney et al., 2024).

Adolescents completed several self-report measures assessing their sleep, including: *the Adolescent Sleep Hygiene Scale (*ASHS; LeBourgeois et al., 2005a), *the Adolescent Sleep Wake Scale* (ASWS; LeBourgeois et al., 2005b), and the PROMIS-PSRI and PROMIS-PSD short forms described above. The ASHS is a 32-item questionnaire that assesses sleep hygiene behaviors over the past month. A total score is derived from the average of all subscale scores (physiological, behavioral arousal, cognitive/emotional, sleep environment, sleep stability, substance use, daytime sleep, and bedtime routine), where higher scores indicate more effective sleep hygiene practices. Prior research has yielded good internal consistency for the total scale (α = .84; Storfer-Isser et al., 2013).

Finally, the ASWS is a 28-item questionnaire that assess sleep quality among youth. Items assess going to bed, falling asleep, maintaining sleep, reinitiating sleep, and returning to wakefulness. Like the ASHS, subscale means (Going to Bed, Falling Asleep and Reinitiating Sleep, Returning to Wakefulness) are averaged to create a total score where higher scores indicate better functioning with respect to sleep. Prior research indicates internal consistency for the overall scale is high (α = .88) and the ASWS has high convergent validity with other gold standard sleep scales (Huber et al., 2020).

#### Interview Measures

Adolescents completed additional interview measures at a baseline study visit. Research personnel administered the *Lifetime Suicide Attempt Self-Injury Interview* (L-SASII; formerly the Lifetime Parasuicide Count; Linehan & Comtois, 1996) to characterize adolescents’ cumulative experiences with suicidal and non-suicidal self-inflicted injury, and the *Childhood Interview for Borderline Personality Disorder* (CI-BPD; Zanarini, 2003) to confirm eligibility. This semi-structured interview assesses all nine BPD criteria from the Diagnostic and Statistical Manual of Mental Disorders (APA, 2022). The interviewer rates each feature as 0 (*absent*), 1 (*probably present*), or 2 (*definitely present*). Prior research indicates items yield good internal consistency (α = .80), high interrater reliability (*k* = .89), and high convergent validity with other BPD measures (Sharp et al., 2012). Finally, youth participants were administered the University of Washington Risk Assessment Protocol at each study visit (Linehan et al., 2012) to aid with risk management (see Ward-Ciesielski & Wilks, 2020).

#### Ecological Momentary Assessment

EMA involves repeated assessment of participants’ experiences in their daily lives. EMA offers greater temporal granularity and ecological validity than traditional longitudinal methods, and helps characterize how experiences of interest may fluctuate or remain stable across short-time intervals within and between persons. Adolescent participants were sent text message prompts to their smartphones (or an email address when no alternative was available) with links to complete online surveys five times per day for two periods throughout the study (10 days pre-intervention and 10-days following an intervention practice period). Survey windows were scheduled based on self-reported wake times, bedtimes, and school schedules for weekdays and weekends. The first prompt (i.e., morning survey) was sent 1 hour before the adolescent’s usual wake time and assessed: sleep (e.g., time participant got into bed last night, time participant tried to fall asleep, duration (minutes) to fall asleep, how often they woke up after sleep onset, awakening duration (minutes), time participant woke up, time participant planned to wake up, time participant left bed, and a rating of sleep quality), momentary mood (positive and negative affective states), wish to live/die, and situational questions (e.g., location, who participants were with). The next three survey prompts were sent semi-randomly within three windows throughout the day (lunchtime, later afternoon [after school], and evening). These prompts assessed momentary mood, momentary wish to live/die, current situation/setting, as well as BPD symptoms and self-harm behaviors *since* their last completed assessment^2^. The last prompt was sent approximately 1 hour before the teen’s usual bedtime. During the pre-intervention period, the assessment included questions about napping, caffeine, momentary mood, cognitive, and situational prompts. Please refer to the EMA protocol supplement for further details on EMA items. Each prompt was spaced at least 1 hour apart. Participants were instructed to complete the surveys as soon as possible. Each survey expired at least 1 hour before the next arrived.

#### EEG

Participants were provided a ZMax wireless EEG headband to record their sleep at home (Hypnodyne Corp., Sofia, Bulgaria; Esfahani et al., 2023). This commercially available ambulatory EEG system was developed for recording physiological sleep data that has been validated against expert-scored polysomnography (PSG) for macro- and micro-structural sleep features (Esfahani et al., 2023; Jafarzadeh Esfahani et al., 2023). The Zmax device acquires EEG activity digitized at 256Hz via two bipolar frontal Ag/AgCL hydrogel electrodes referenced to a frontal ground (Fpz). The two channels correspond to F7-Fpz (left frontal) and F8-Fpz (right frontal) on a traditional 10-20 EEG montage (Klem et al., 1999). The bipolar configuration provides bilateral prefrontal coverage without a mastoid reference channel which facilitates at-home use but differs from conventional sleep PSG montages (Esfahani et al., 2023).

All EEG preprocessing steps were performed using standard functions in MATLAB (Mathworks, 2024). To attenuate slow signal drift throughout each night of sleep, EEG data were detrended by subtracting a 10s moving average from each channel (Widmann et al., 2015). We also performed a discrete wavelet denoising procedure (Daubechies db4, decomposition level 10) to mitigate multiscale effects of physiological artifacts and signal noise on later sleep scoring (Al-Qazzaz et al., 2015; Mallat, 1989; Mamun et al., 2013).

The Dreamento toolbox (Jafarzadeh Esfahani et al., 2023) was used for sleep-staging. This open-source Python toolbox uses a custom-built classifier (DreamentoScorer) to classify epochs of EEG data into the canonical sleep stages (i.e., Wake, N1, N2, N3, REM). and the Yet Another Spindle Algorithm (YASA; Vallat & Walker, 2021) to detect sleep-related events. Automatic eye movement detection and automatic slow oscillation plus spindle detection was enabled with outlier removal for each night of sleep EEG (Vallat & Walker, 2021). The standard Dreamento pipeline epoched the EEG into 30s segments and applied a 0.3 - 30 Hz bandpass filter to both channels. Dreamento then performed an ±3 epoch-wise feature-extraction procedure (e.g., signal statistics, spectral properties, non-linear dynamics) which was normalized and classified data into sleep stages using Light Gradient Boosting Machine (LightGBM). Once classified, Dreamento passed the scored sleep hypnograms to YASA’S sleep statistics analysis (Vallat & Walker, 2021) to derive macroscale sleep metrics for each night. These include time in bed (time from the beginning to the end of the EEG recording), sleep period time (time from first sleep epoch to final sleep epoch), total sleep time (sleep period time minus wakings [in minutes]), wake after sleep onset (number of wake epochs [in minutes] after sleep onset), sleep latency onset (time from the start of the recording to first sleep epoch), and sleep maintenance efficiency (total sleep time **÷** sleep period time × 100).

To select files for inclusion, we visually inspected all EEG tracings and spectrograms. Using a three-member consensus approach, we selected recordings to retain for sleep scoring that were of sufficiently high quality and plausibility when compared to participants’ EMA reports and a participant-maintained paper-and-pencil log of headband recording times.

### Procedure

After consent procedures, adolescents were scheduled for a brief virtual visit with research personnel to complete the UWRAP, SIQ-Jr., CI-BPD, and the L-SASII. At the end of the visit, adolescents and their caregivers were sent their baseline survey packets, scheduled to pick-up their sleep monitoring headband, and youth were oriented to completing EMA surveys for a period of 10 days. The study team confirmed all participants completed their baseline survey packets prior to receiving their EEG headbands. Adolescent EMA began the morning following participants’ first night of EEG recording. At study outset, youth made best attempts to record their sleep at home via EEG for up to 10 nights before and after the sleep intervention; however, at approximately one third through the study, we shifted our protocol such that participants attempted to collect up to 7 nights of EEG data pre- and post-intervention^3^.

Following the baseline EEG/EMA period, participants returned their devices and were scheduled for their intervention session. This 1.5-to-2-hour virtual visit was completed with the adolescent, their caregiver, and a trained clinician^4^. During this visit, participants engaged in several key activities from the TSC-Youth intervention: (1) a functional analysis, (2) psychoeducation, (3) goal setting, (4) creating a personalized rise-up routine, (5) and wind-down routine, and (6) reviewing stimulus control guidelines for high-quality sleep. The clinician collaboratively conducted a functional analysis on the adolescent’s sleep-related habits with their caregiver’s input. We documented what the adolescent does, thinks, and feels at bedtime, in the night (should they wake up), on waking, and noted daytime behaviors that could impact sleep and energy levels (e.g., napping, caffeine use, exercise, meal-times, etc.). Following the functional analysis, participants watched a 20-min psychoeducational video created by the research team presenting key content from the TSC-Youth intervention on the importance of sleep, common pitfalls that can disrupt sleep quality (e.g., waking at inconsistent times during the week vs. the weekend, exercising or eating a large meal too close to bed, exposure to screens and bright lights within 1-2 hours of trying to initiate sleep, worrying in bed), and evidence-based sleep hygiene practices. Following the video, participants revisited their functional analysis to add any relevant behaviors they may have missed, and were guided through a companion handout on improving their sleep.

The clinician next worked with the adolescent to set a series of concrete and specific goals for daytime and nighttime behavior for them to enact during the remainder of the study (e.g., choosing a feasible bedtime and wake-time over the subsequent weeks to incrementally shift participants’ sleep schedules, limiting caffeine use to the morning hours and swapping in caffeine-free alternatives for afternoons and evenings, scheduling time to worry/plan earlier in the day). Clinicians engaged caregivers throughout the intervention to enhance familial support for creating an environment and routine conducive to high-quality sleep. Participants were provided a morning checklist of activities to perform upon waking to enhance alertness (RISE UP routine), and generated a personalized wind-down routine. This routine lists activities to perform in the hour(s) before bed that are conducive to low-light and relaxation, as well as activities to avoid. Finally, the clinician reviewed a stimulus control handout with the family. At the close of the session, youth and caregiver participants were emailed copies of the intervention handouts and notes from the visit and asked to practice their new routines for the remainder of the study. Participants were sent a text message from the study team approximately 1-week post-intervention to check in about their home practice.

Approximately 4 weeks post-intervention, participants repeated their 10-day EMA assessment and 7- to 10-day EEG protocols. Caregivers and youth were sent their second, post-intervention, survey packet upon completion of the second EMA period. A final survey packet was distributed one-month later.

#### Analytic Plan

We used MATLAB’s [fitlme] function (Mathworks, 2024) to compare survey packets between study phases with linear mixed effects models (LMEs). We specified that each LME model use random intercepts and directional one-tailed tests corresponding to improvements in BPD symptoms or sleep. To correct for multiple comparisons, surveys were grouped into youth-report sleep-related assessments, youth-reported BPD-related questionnaires, and caregiver-reports on youth sleep (i.e., family-wise groupings). We accounted for multiple comparisons using Benjamini–Hochberg false discovery rate (FDR) correction based on the number of tests in each group. Youth report on the PROMIS-PSRI and PROMIS-PSD were primary outcomes for this study, whereas other sleep and BPD feature measures reflect secondary outcomes.

To prepare for EEG analyses, we preprocessed and sleep-scored each night of EEG data. We first calculated macroscale sleep metrics to quantify average sleep quality across the sample. To test our hypothesis that participants’ sleep quality would improve, we used a LME model with random intercepts to compare time in bed, sleep period time, total sleep time, wake after sleep onset, sleep onset latency, and sleep maintenance efficiency.

Analyses based upon EMA data were conducted using MPlus version 9.1 (Muthén & Muthén, 1998-2026). A two-level, random effects model using robust maximum likelihood estimation (MLR estimator) was specified in which the sleep-relevant construct of interest was estimated at the between-and within-person level. The MLR estimator is appropriate for a variety of dependent variable types and non-normal data, and uses full information maximum likelihood (FIML) to handle missing data. FIML estimation avoids the limitations of dropping or imputing missing data, and yields unbiased estimates for data missing completely at random or missing at random (Xiao & Bulut, 2020). At the within-person level, this allowed us to examine fluctuations relative to the person’s own observed mean, and, at the between-person level, fluctuations relative to the overall sample mean. At the within-person level, the sleep variable was regressed on a binary variable for time (0 = pre-intervention, 1 = post-intervention) and modeled with a random slope. The sleep parameter was also regressed onto a binary variable indicating whether the participant attended school (1) or was home (0, school holiday or weekend) on the following day, as adolescents often exhibit distinct patterns of sleep preceding days with and without school attendance. Thus, all models controlled for the within-person effect of being at home (versus out of the home) on prior night sleep parameters. The random slope parameter was allowed to covary with the sleep parameter at the between-person level.

For BPD symptoms, latent variables were estimated at the within and between person levels with categorical (binary) specified indicators for eight items tapping BPD symptomology since the last survey. Variables tapping wish to live and wish to die were modeled together, allowing them to covary with each other at the within-and between-person level, given their high covariance. The same approach was used for modeling negative affect and positive affect simultaneously, in a separate model.

## Results

Demographic characteristics for the sample are summarized in Table 1 with baseline clinical characteristics of adolescent participants reported in Table 2. Results from LME models examining changes in self- and caregiver-report scales pre- to post-intervention are presented in Table 3. Notably, all adolescent and caregiver sleep-focused measures significantly improved in the post-intervention and 1-month follow-up period relative to pre-intervention baseline. Although BPFSC-11 scores did not meaningfully shift from pre- to post-intervention, many other indices of core BPD features significantly improved (e.g., scores on the DERS-SF, SCIM, and most subscales of the UPPS-P).

**Table 1.**
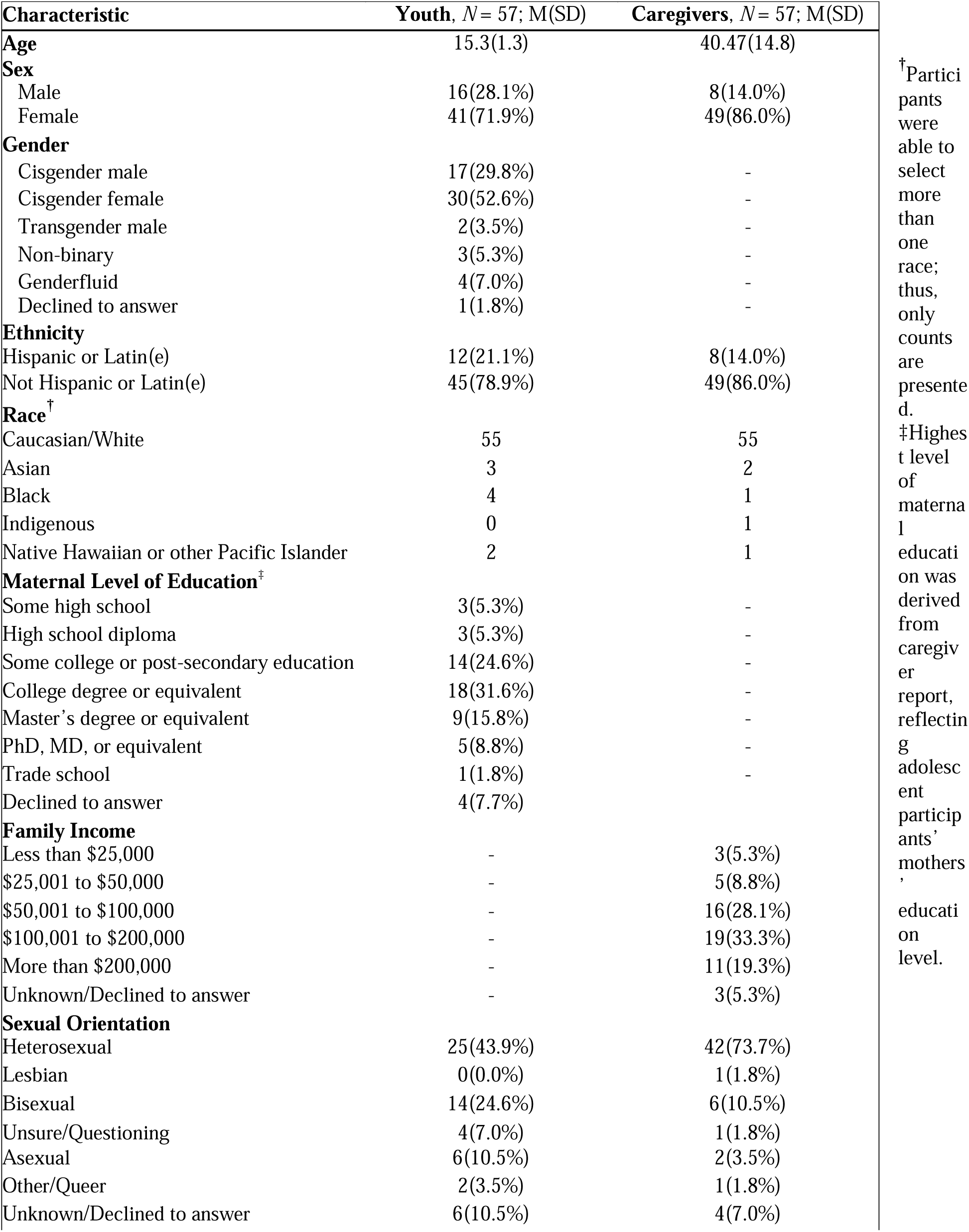
Detailed Baseline Demographic Variables.

**Table 2.**
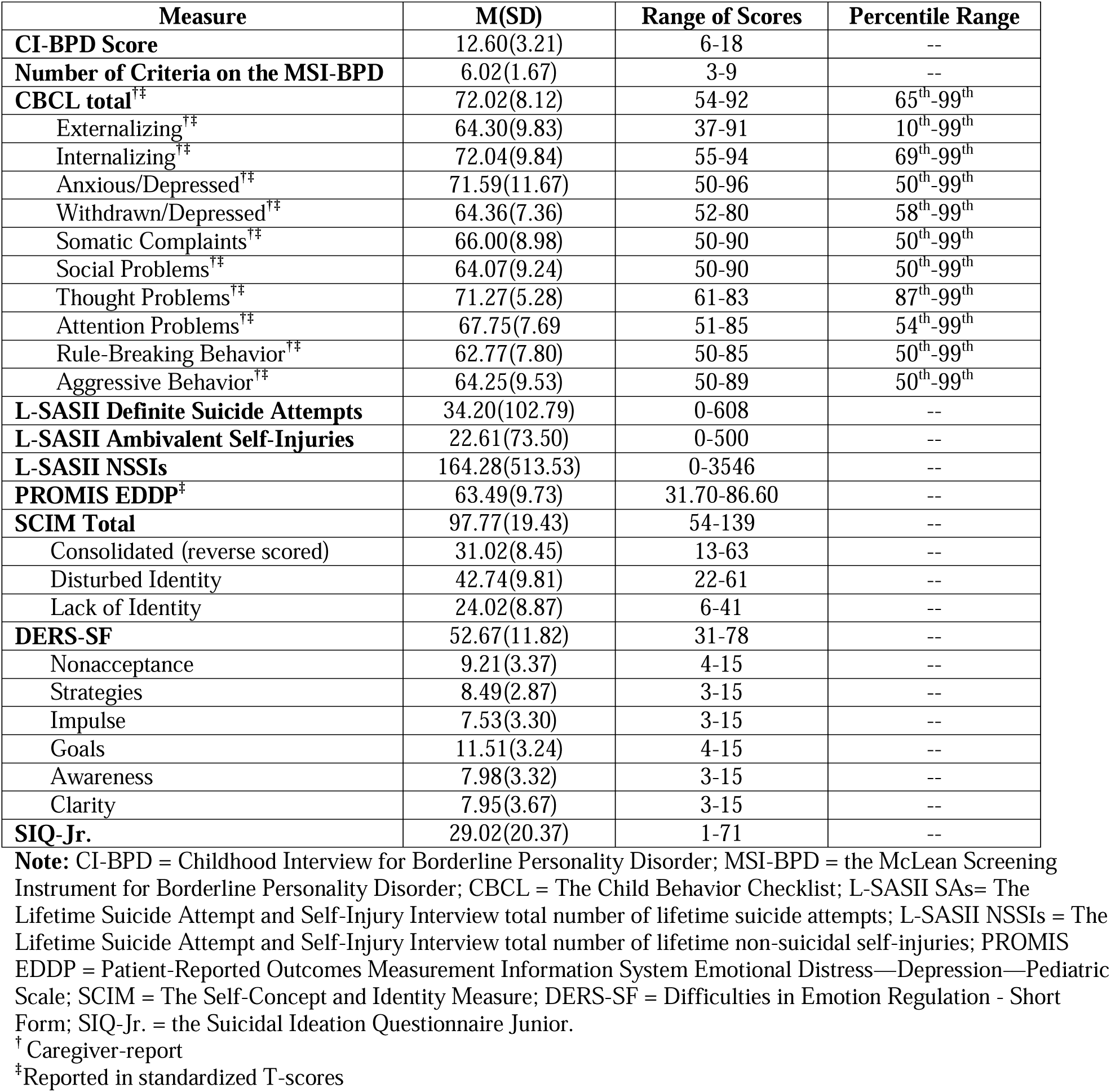
Youth baseline clinical characteristics.

| Measure | M(SD) | Range of Scores | Percentile Range |
| --- | --- | --- | --- |
| **CI-BPD Score** | 12.60(3.21) | 6-18 | -- |
| **Number of Criteria on the MSI-BPD** | 6.02(1.67) | 3-9 | -- |
| **CBCL total<sup>†‡</sup>** | 72.02(8.12) | 54-92 | 65<sup>th</sup>-99<sup>th</sup> |
| Externalizing<sup>†‡</sup> | 64.30(9.83) | 37-91 | 10<sup>th</sup>-99<sup>th</sup> |
| Internalizing<sup>†‡</sup> | 72.04(9.84) | 55-94 | 69<sup>th</sup>-99<sup>th</sup> |
| Anxious/Depressed<sup>†‡</sup> | 71.59(11.67) | 50-96 | 50<sup>th</sup>-99<sup>th</sup> |
| Withdrawn/Depressed<sup>†‡</sup> | 64.36(7.36) | 52-80 | 58<sup>th</sup>-99<sup>th</sup> |
| Somatic Complaints<sup>†‡</sup> | 66.00(8.98) | 50-90 | 50<sup>th</sup>-99<sup>th</sup> |
| Social Problems<sup>†‡</sup> | 64.07(9.24) | 50-90 | 50<sup>th</sup>-99<sup>th</sup> |
| Thought Problems<sup>†‡</sup> | 71.27(5.28) | 61-83 | 87<sup>th</sup>-99<sup>th</sup> |
| Attention Problems<sup>†‡</sup> | 67.75(7.69) | 51-85 | 54<sup>th</sup>-99<sup>th</sup> |
| Rule-Breaking Behavior<sup>†‡</sup> | 62.77(7.80) | 50-85 | 50<sup>th</sup>-99<sup>th</sup> |
| Aggressive Behavior<sup>†‡</sup> | 64.25(9.53) | 50-89 | 50<sup>th</sup>-99<sup>th</sup> |
| **L-SASII Definite Suicide Attempts** | 34.20(102.79) | 0-608 | -- |
| **L-SASII Ambivalent Self-Injuries** | 22.61(73.50) | 0-500 | -- |
| **L-SASII NSSIs** | 164.28(513.53) | 0-3546 | -- |
| **PROMIS EDDP<sup>‡</sup>** | 63.49(9.73) | 31.70-86.60 | -- |
| **SCIM Total** | 97.77(19.43) | 54-139 | -- |
| Consolidated (reverse scored) | 31.02(8.45) | 13-63 | -- |
| Disturbed Identity | 42.74(9.81) | 22-61 | -- |
| Lack of Identity | 24.02(8.87) | 6-41 | -- |
| **DERS-SF** | 52.67(11.82) | 31-78 | -- |
| Nonacceptance | 9.21(3.37) | 4-15 | -- |
| Strategies | 8.49(2.87) | 3-15 | -- |
| Impulse | 7.53(3.30) | 3-15 | -- |
| Goals | 11.51(3.24) | 4-15 | -- |
| Awareness | 7.98(3.32) | 3-15 | -- |
| Clarity | 7.95(3.67) | 3-15 | -- |
| **SIQ-Jr.** | 29.02(20.37) | 1-71 | -- |

**Note:** CI-BPD = Childhood Interview for Borderline Personality Disorder; MSI-BPD = the McLean Screening Instrument for Borderline Personality Disorder; CBCL = The Child Behavior Checklist; L-SASII SAs= The Lifetime Suicide Attempt and Self-Injury Interview total number of lifetime suicide attempts; L-SASII NSSIs = The Lifetime Suicide Attempt and Self-Injury Interview total number of lifetime non-suicidal self-injuries; PROMIS EDDP = Patient-Reported Outcomes Measurement Information System Emotional Distress—Depression—Pediatric Scale; SCIM = The Self-Concept and Identity Measure; DERS-SF = Difficulties in Emotion Regulation - Short Form; SIQ-Jr. = the Suicidal Ideation Questionnaire Junior.
<sup>†</sup> Caregiver-report
<sup>‡</sup>Reported in standardized T-scores

**Table 3.** Intervention effects on self- and informant-report scales.

| Measure | Baseline | Post | 1 Month | Pre to post‡ |  |  |  | Pre to 1-Month follow-up‡ |  |  |  |
| --- | --- | --- | --- | --- | --- | --- | --- | --- | --- | --- | --- |
|  | *M (SD)* | *M (SD)* | *M (SD)* | *b* | *t* | *p* | *d<sub>z</sub>* | *b* | *t* | *p* | *d<sub>z</sub>* |
| **ASHS†** | 4.03(0.60) | 4.54(0.54) | 4.6(0.62) | 0.52 | 7.06 | < .001 | 0.91 | 0.56 | 7.65 | < .001 | 0.88 |
| **ASWS†** | 2.94(0.74) | 3.60(0.74) | 3.64(0.75) | 0.66 | 5.83 | < .001 | 0.73 | 0.69 | 6.06 | < .001 | 0.68 |
| **PROMIS PSRI†** | 66.86(7.54) | 59.36(8.86) | 57.31(7.58) | -7.50 | -6.30 | < .001 | -0.80 | -9.50 | -7.93 | < .001 | -1.00 |
| **PROMIS PSD†** | 65.68(6.35) | 57.80(6.03) | 56.61(5.89) | -7.89 | -9.15 | < .001 | -1.19 | -8.97 | -10.35 | < .001 | -1.18 |
| **BPFSC-11†** | 33.72(8.30) | 33.61(8.70) | -- | -0.11 | -0.11 | .46 | -0.02 | -- | -- | -- | -- |
| **DERs-SF†** | 52.67(11.82) | 48.25(13.99) | 44.59(12.99) | -4.33 | -2.67 | .004 | -0.34 | -7.95 | -4.89 | < .001 | -0.57 |
| **SCIM†** | 97.77(19.43) | 90.39(23.31) | 88.13(23.21) | -7.39 | -2.97 | .002 | -0.36 | -9.61 | -3.84 | < .001 | -0.46 |
| **UPPS-P†** |  |  |  |  |  |  |  |  |  |  |  |
| Negative Urgency | 2.99(0.64) | 2.75(0.81) | 2.73(0.82) | -0.25 | -2.85 | .01 | -0.37 | -0.25 | -2.94 | .01 | -0.36 |
| Lack of Premeditation | 2.30(0.64) | 2.14(0.65) | 2.13(0.60) | -0.17 | -2.30 | .01 | -0.31 | -0.17 | -2.33 | .02 | -0.28 |
| Lack of Perseverance | 2.02(0.50) | 1.88(0.53) | 1.98(0.41) | -0.14 | -2.26 | .03 | -0.32 | -0.04 | -0.63 | .30 | -0.07 |
| Sensation seeking | 2.58(0.68) | 2.65(0.65) | 2.60(0.69) | 0.08 | 1.12 | .30 | 0.14 | 0.04 | 0.52 | .60 | 0.06 |
| Positive Urgency | 1.39(0.76) | 2.23(0.88) | 2.28(0.79) | -0.16 | -1.94 | .05 | -0.22 | -0.10 | -1.20 | .17 | -0.15 |
| **Caregiver PROMIS PSRI†** | 67.60(8.56) | 61.27(9.32) | 62.083(8.09) | -6.37 | -5.04 | < .001 | -0.64 | -5.57 | -4.36 | < .001 | -0.57 |
| **Caregiver PROMIS PSD†** | 65.10(9.31) | 58.95(8.27) | 58.44(9.32) | -6.20 | -6.04 | < .001 | -0.77 | -6.23 | -6.37 | < .001 | -0.76 |

**Note:** ASHS = Adolescent Sleep Hygiene Scale; ASWS = Adolescent Sleep Wake Scale; PROMIS PSRI = Patient-Reported Outcomes Measurement Information System Pediatric Sleep-related Impairment Short-Form; PROMIS PSD = PROMIS Pediatric Sleep Disturbances Short-Form; BPFSC-11 = Borderline Personality Features Scale for Children-11; DERs-SF = the Difficulties in Emotion Regulation Short-Form total; SCIM = Self-Concept and Identity Measure total; UPPS-P = the Urgency, Premeditation (lack of), Perseverance (lack of), Sensation Seeking, Positive Urgency, Impulsive Behavior Scale. Cohen's $d_z$ values reflect magnitude of paired change scores pre- to post-intervention. †p-values are FDR corrected. ‡ Tests were one-tailed in the direction of improvement.

### EEG Results

We received a total of 783 EEG recordings from participants (405 pre-intervention, 367 post-intervention; 11 unmatched to phase and excluded). Unavoidable and frequent failures of the Zmax device in-home, recurrent data corruption, and significant noise contamination among individual EEG files led us to exclude a substantial portion of data from later analyses. Overall, we retained 27 participants with 170 nights of sleep EEG for analysis (90 pre-intervention, 80 post-intervention).

During the pre-intervention phase, mean time in bed was 8.32 hours (*SD* = 2.08 hours, range = 3.38 – 14.05 hours). Average sleep period time across all nights of 8.03 hours (*SD* = 2.08 hours, range = 3.13 – 13.88 hours) with average total sleep time of 6.90 hours (*SD* = 1.85 hours, range = 2.36 – 10.88 hours). Mean sleep onset latency across all pre-intervention nights was 13.29 minutes (*SD* = 26.95 minutes, range = 0.00^5^ – 238.00 minutes) and we found that average time of wake after sleep onset was 1.11 hours (*SD* = 1.05 hours, range = 0.133 – 5.75 hours). At the pre-intervention phase, mean sleep maintenance efficiency was 86.37% (*SD* = 11.10%, range = 41.39 – 98.56%) across all nights.

During the post intervention phase, average time in bed was 8.80 hours (*SD* = 1.79 hours, range = 3.72 – 18.11). Average sleep period time post-intervention was 8.53 hours (*SD* = 1.73 hours, range = 3.68 – 16.89) and mean total sleep time was 7.54 hours (*SD* = 1.48 hours, range = 3.37 – 15.13). Average sleep onset latency post-intervention was 13.25 minutes (*SD* = 15.20 minutes, range = 0* - 73.00 minutes) and time spent awake after sleep onset was 1.00 hour (*SD* = 0.75 hours, range = 0.03 – 3.63). Across all post intervention nights, we found an average sleep maintenance efficiency of 88.82% (*SD* = 7.61%, range = 55.79 -99.57%).

We compared macrostructural sleep statistics between pre- and post-intervention using a LME with random intercepts (one-tailed; Cohen’s *d_z_* reflects magnitude of paired change) and found time in bed increased significantly in the post-intervention phase (*t*(168) = 2.41, *p_FDR_ =* 0.03, *B =* 0.62, 95% CI = 0.11 - 1.12, Cohen’s *d_z_* = 0.46). Likewise, sleep period time significantly increased (*t*(168) = 2.62, *p_FDR_*= 0.01, *B* = 0.66, 95% CI = 0.16 – 1.15, Cohen’s *d_z_* = 0.54) along with total sleep time (*t*(168) = 3.06, *p_FDR_* = 0.01, *B* = 0.71, 95% CI = 0.25 – 1.16, Cohen’s *d_z_* = 0.59). Although the other macroscale sleep metrics were not significantly different between phases, sleep onset latency, wake after sleep onset, and sleep maintenance efficiency all trended in the expected direction post-intervention (see Figure 2).

**Figure 2.**
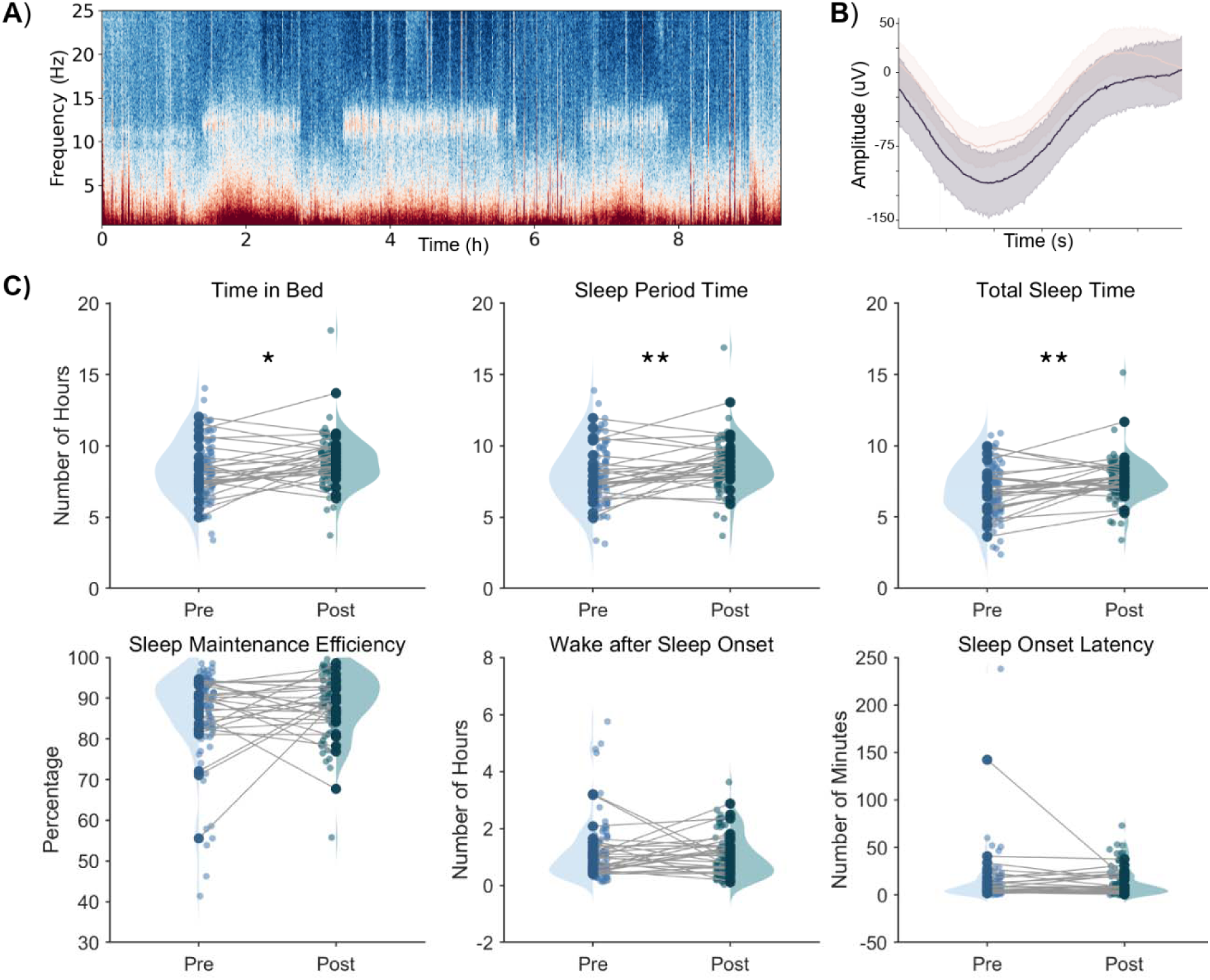
Example EEG spectrogram, average slow-wave, and all sleep statistical analyses. **A)** Example spectrogram showing EEG frequency power over time for a single night of sleep (warmer colors = higher power). **B)** Average slow wave for a single night of sleep during N2 (pink) and N3 (grey). **C)** Raincloud plots showing sleep statistics pre-intervention (*n* = 90) and post-intervention (*n* = 80). Pale dots represent single nights of sleep; dark dots represent subject averages. ***Note***: * = *p* < 0.05; ** = *p* < 0.01, FDR corrected. One outlier night removed for visualization in **C)** but included in analyses.

### EMA Results

Participants completed a total of 4,179 assessments (overall adherence = 74.6%, range = 43% - 98%). On average, participants reported an earlier clock time for when they fell asleep (*b* = -47.47, *se* = 11.39, *p* < .001) and when they woke up (*b* = -24.94, *se* = 8.18, *p* = .002) after the intervention, relative to pre-intervention data. Participants also reported significantly greater total sleep time (in minutes) following the intervention (*b* = 35.64, *se* = 11.11, *p* = .001). Participants reported fewer total awakenings after sleep onset in the post-intervention EMA period (*b* = -0.25, *se* = -0.11, *p* = .02), although there was no effect on total number of minutes awake after sleep onset (*b* = -4.05, *se* = 4.07, *p* = .32). There was weak evidence of improvement in self-reported sleep quality (*b* = 0.16, *se* = 0.08, *p* = .06), and no effect on number of minutes to fall asleep after getting into bed (*b* = -4.88, *se* = 3.59, *p* = .17). Participants endorsed taking fewer sleep-related medications post-treatment (*b* = -0.13, *se* = 0.02, *p* = < 0.0001). Post-treatment, all but one participant endorsed using TSC skills and routines on EMA^6^ (*M* = 7.05 days, *SD* = 2.45 days out of the 10-day assessment window). Participants reportedly used a wake routine, on average, 67% of EMA days; timed light exposure on 62% of days; stuck to sleep schedule on 46% of days; used wind down routine 57% of days; and used sleep health practices 75% of days with available data. Of the days where sleep health practices were endorsed, participants reported turning off screens and lights 75% of days, keeping electronics out of their bedroom 45% of days, avoided eating near bedtime 75% of days, and engaging in exercise 50% of surveyed days.

With respect to improvement in psychopathology and related constructs, BPD symptoms reported during EMA were significantly reduced following the intervention (*b* = -1.57, *se* = 0.34, *p* < .001). To inform interpretation of null findings with the BPFSC-11, which primarily taps interpersonal dysfunction, a supplementary analysis examining frequency of reported arguments during EMA (one indicator of the BPD latent factor) specifically changed post-intervention; arguments did, in fact, decrease after treatment (*b* = -0.80, *se* = 0.27, *p* = .003). There was also significant improvement in negative affect (*b* = -3.95, *se* = 1.84, *p* = .03) but not positive affect (*b* = 0.91, *se* = 1.53, *p* = .55) when modeled concurrently. There were no significant intervention effects on concurrently modeled wish to live (*b* = 1.93, *se* = 1.72, *p* = .26) or wish to die (*b* = - 1.03, *se* = 1.51, *p* = .49). Detailed EMA results are reported in supplemental tables.

## Discussion

Results from convergent methods across multiple-levels of analysis and multiple reporters indicate that adolescents’ sleep and BPD features shifted following a single-session sleep-focused intervention. Not only did many indices of sleep quantity and quality improve across both objective (EEG) and subjective assessments (EMA and self- and caregiver-report), but multiple indices of BPD symptoms also improved significantly after three to four weeks of at-home sleep routine practice. Although some metrics failed to shift (e.g., BPFSC-11 scores did not significantly change, there was no effect on wish to live/die, number of minutes awake after sleep onset or number of minutes to fall asleep after getting into bed), many important indices robustly improved after treatment. During EMA, youth reported earlier bedtimes (∼47 minutes earlier) and waketimes (∼25 minutes earlier), significantly more sleep (∼36 minutes), and fewer awakenings after the intervention. Although only a fraction of participants (*n* = 9) reported engaging in TSC skills and routine practice every day of the EMA follow-up period, almost all participants did endorse some skills practice one-month following the single-session intervention. Further, *every* sleep-related caregiver and self-report questionnaire not only improved at post-intervention, but was maintained at 1-month follow-up relative to baseline (capturing sleep hygiene and practices, sleep disturbances, and sleep-related impairment). Youth endorsed taking less sleep-related medications in the post-intervention period, indicating improvements are more likely to be attributable to behavioral changes. BPD symptoms reported via EMA were significantly reduced, as was negative affect in the post-intervention period. These findings were complemented by improvements in measures of identity-related functioning, emotion dysregulation and most, but not all facets of impulsivity (negative urgency, lack of premeditation, lack of perseverance, and positive urgency scores^7^).

Results from the present trial are promising for several reasons. First, adolescents in our sample were considerably impaired, yet many positive changes were apparent following our low-intensity intervention. Although participants were eligible with as few as three BPD symptoms, the average number of criteria endorsed in our sample was greater than six— surpassing the diagnostic threshold of five—with some participants reporting all nine symptoms. Other psychopathology measures used to characterize the sample also point to marked difficulties at baseline (see Table 2; 80.7% of participants endorsed a history of self-injury, with 56.1% having made a suicide attempt). Consistent with prior research demonstrating sleep disruptions among BPD samples (Jenkins et al., 2022), participants’ baseline EEG duration was often irregular, with some adolescents experiencing very short nights of sleep (minimum: 3.38 hours) and others sleeping excessively (maximum: 14.05 hours). Many experienced significant sleep delays (taking hours to fall asleep and/or initiating sleep well past midnight) and excessive nighttime awakening (5 hours as reported on EMA). Notably, participants were not recruited immediately following hospitalization or major crisis. This both highlights the great need for intervention among at-risk adolescents as young as 13, and helps guard against regression to the mean that is common following a period of acute crisis. Although many clinicians may be reluctant to focus on sleep with this population due to concerns about treatment adherence, motivation, or out of a desire to focus more directly on treating the features of BPD, results of this trial support sleep as a meaningful intervention target. Regulating sleep may have a positive impact on youth symptoms in day-to-day life, and on validated measures of key BPD features.

Second, effects were observed after a single ∼1.5-hour treatment session. Content taught in the TSC-Youth intervention is designed as an accessible, straightforward, and easy-to-disseminate approach to treat a wide range of sleep and circadian problems impacting adolescents (e.g., eveningness circadian preference, daytime sleepiness, discrepancy in sleep and wake times on weekends vs. weekdays; Harvey et al., 2018; Harvey & Buysee, 2017). Unlike frontline treatments for BPD such as DBT, mentalization-based therapy, and transference-focused psychotherapy which typically last 6 to 18+ months and require a high-degree of training for adherent administration, the TSC-Youth intervention is brief, and can be delivered by a wider array of service providers. BPD is a serious mental illness that is highly stigmatized and often considered difficult to treat (Masland et al., 2023). Although affected individuals are overrepresented among those seeking medical and psychiatric services (Pascual et al., 2007; Sansone et al., 2011), many go without effective care (Masland et al., 2023). The field is in urgent need of “low-hanging fruit” interventions that may serve as secondary or tertiary prevention to: help reduce the impact of BPD early in development before symptoms become more entrenched, make symptoms more manageable, and potentially increase readiness for other forms of care (regulated and sufficient sleep may enhance memory and attentional processes vital for psychotherapy; Mander et al., 2011).

Third, our study design prioritized ecological validity, as EEG and EMA were each recorded in the everyday home environment. Although much of our EEG data were removed from analyses, changes were still apparent. Attaining ambulatory sleep data in adolescents’ home environments is almost certainly better capturing their day-to-day sleep than data collected in a laboratory environment. Although BPFSC-11 scores did not change pre-to post-intervention, more proximally assessed symptoms improved in daily life. The BPSC-11 largely assesses negativity in interpersonal relationships. It may be that participants’ negative perceptions of their relationships are more durable (less responsive to the intervention), or simply slower to change compared with other indicators of risk. Even if participants may have begun handling interpersonal situations more effectively, interaction partners may continue to be skeptical or rejecting for a longer period of time. When we examined arguing as assessed via EMA, we did see fewer overall incidents in the post-intervention period. Thus, although participants’ ratings on the BPFSC-11 did not significantly change, their proximal, day-to-day experience of symptoms shifted.

As with all research, our study has notable limitations. First, although we made every effort to recruit families from diverse backgrounds, our sample was predominantly female, and the demographics of our sample roughly match those of the state from which participants were recruited. It is unclear how well findings will generalize males and to non-majority white samples. We also experienced significant problems collecting full nights of ambulatory EEG sleep data. However, EEG data that were analyzed are high-quality and do align with other sources of information/results from the study. EMA assessments of suicidal ideation and self-injurious urges and behaviors were not endorsed with sufficient frequency or variability to examine in multi-level models. Thus, further work is needed to evaluate this important outcome.

Importantly, we did not include a control group in the current trial due to funding and time constraints. Future randomized controlled trial(s) are essential for ruling out regression to mean, spontaneous recovery, placebo and expectancy effects, and to ensure benefits are indeed attributable to the sleep intervention rather than potential concurrent treatment or other factors. Finally, our study only followed participants for approximately 2 months post-intervention (post-intervention assessment took place after 1 month of practice followed by an additional 1-month follow-up survey battery that did not include interview measures or the BPFC-11). The durability of intervention effects needs to be more comprehensively assessed at longer time intervals. Results from meta-analytic studies of single-session randomized clinical trials targeting psychiatric problems have found effect sizes are typically in the small to medium range (Schleider & Weisz, 2017). Unsurprisingly, effect sizes were smaller for trials included an active control condition (as compared to no treatment) and among trials that included longer-follow up periods (more than 13 weeks). They also found that effect sizes were larger for cognitive-behavioral approaches and for child samples relative to teens. Given this pattern, we can assume that the large to medium effect sizes observed in the present study may be smaller if we had included an active control condition or included a longer follow-up period.

The present study represents a first step in investigating the a sleep intervention for youth with BPD features. Future trials should incorporate an assessment only control group, explicitly test feasibility, and examine potential incremental benefits of the full multi-session TSC-Youth intervention. Should efficacy become better established, future work may explore whether a stepped care approach should be recommended (e.g., sleep intervention prior to gold-standard BPD treatment models), and assess whether frontloading attention on sleep regulation may enrich established interventions. We elected to include a caregiver in the intervention, as parental involvement is often beneficial for this population (see Bosworth et al., 2024 for a review). Yet future work should investigate adolescent-only treatment for cases when a caretaker may not be available. Additional research is needed to assess intervention delivery in diverse treatment settings (e.g., primary care, psychiatry), and by a range of provider types (e.g., physicians, social workers, or even bachelors-level professionals). Results of the current trial are promising with many findings evincing moderate to large effect sizes. It may be possible that even low-intensity treatments that aid with physiological regulation may yield meaningful benefits to youth vulnerable to BPD.

## Data Availability

All data produced in the present study are available upon reasonable request to the authors

## Footnotes

1 For caregivers without such a consent form available in their records, research assistants sent an email with broad information about the study that gave a clear option to opt out of further communication. These caregivers were contacted further via phone if (1) they expressed interest in response to the email or (2) they did not opt out of further communication within a week of receiving the email.

2 A safety monitoring protocol was in place for self-injurious thoughts and behaviors (see EMA protocol in supplementary material).

3 This change was made for a number of reasons. First, it reduced participant burden, as wearing the device was challenging for some to remember and uncomfortable for some youth to wear. Second, this change significantly simplified our protocol as the Zmax device stores data on micro-SD cards. Each SD card holds up to 8 recordings. Reducing the EEG period to seven nights meant that the study staff could include a test recording and collect the entire period of sleep data without asking participants to change the SD card. Third, scheduling with families became more efficient, as many were available at the same time for pick-up/drop-off week-to-week.

4 The two study clinicians were a clinical psychology Ph.D. candidate supervised by the first author, and the first author herself, who is a licensed psychologist.

5 Sleep onset latency of 0 can occur if participants fall asleep within one epoch of recording start.

6 This participant did not complete any end of day assessments during the post-intervention period, which were the only assessments including items capturing skills use. Thus, it is possible that the whole sample employed skills from the intervention, but we cannot be certain.

7 Sensation seeking on the UPPS-P was unaffected; however, this subscale demonstrated poor internal consistency in the present sample, and items do not necessarily tap psychopathology (e.g., enjoying risk-taking, welcoming exciting experiences/sensations that may be frightening or unconventional, liking to learn to fly an airplane, enjoying the sensation of fast skiing). Changes on lack of perseverance and positive urgency were not maintained at 1-month follow-up.

