## Supplement for "Improved Sleep and Clinical Outcomes Following a Single-Session Intervention among Adolescents with Borderline Personality Disorder Features"

**Supplemental Self- and Caregiver-Report Internal Consistencies in the Present Sample**

Table 1. *Internal Consistencies for self- and caregiver-report scales at each time point of administration in the current sample*

| **Measure** | **Baseline** | **Post-Intervention** | **1-Month Follow-up** |
| --- | --- | --- | --- |
| ***ADOLESCENT-REPORT*** | | | |
| PROMIS-PSD | α = .60 | α = .77 | α = .74 |
| PROMIS PSRI | α = .88 | α = .92 | α = .91 |
| PROMIS EDDP | α = .93 | -- | -- |
| BPFSC-11 | α = .85 | α = .85 | -- |
| DERS-SF Total | α = .85 | α = .86 | α = .86 |
| Nonacceptance | α = .83 | α = .84 | α = .88 |
| Strategies | α = .76 | α = .73 | α = .83 |
| Impulse | α =. 88 | α = .93 | α = .96 |
| Goals | α = .89 | α = .94 | α = .95 |
| Awareness | α = .86 | α = .89 | α = .88 |
| Clarity | α = .90 | α = .89 | α = .81 |
| UPPS-P |  |  |  |
| Negative Urgency | α = .68 | α = .82 | α = .82 |
| Lack of Premeditation | α = .76 | α = .83 | α = .75 |
| Lack of Perseverance | α = .49 | α = .70 | α = .48 |
| Sensation seeking | α = .60 | α = .50 | α = .62 |
| Positive Urgency | α = .74 | α = .83 | α = .80 |
| SCIM Total | α = .78 | α = .84 | α = .86 |
| Consolidated (reverse scored) | α = .64 | α = .65 | α = .77 |
| Disturbed Identity | α = .65 | α = .75 | α = .82 |
| Lack of Identity | α = .82 | α = .83 | α = .87 |
| SIQ Jr. | α = .96 | -- | -- |
| ASHS | α = .87 | α = .85 | α = .85 |
| ASWS | α = .67 | α = .59 | α = .59 |
| ***PARENT-REPORT*** | | | |
| CBCL Total | α = .93 | -- | -- |
| Externalizing | α = .90 | -- | -- |
| Internalizing | α = .89 | -- | -- |
| Anxious/Depressed | α = .89 | -- | -- |
| Withdrawn/Depressed | α = .64 | -- | -- |
| Somatic Complaints | α = .75 | -- | -- |
| Social Problems | α = .80 | -- | -- |
| Thought Problems | α = .59 | -- | -- |
| Attention Problems | α = .73 | -- | -- |
| Rule-Breaking Behavior | α = .81 | -- | -- |
| Aggressive Behavior | α = .88 | -- | -- |
| Parent PROMIS PSD | α = .80 | α = .80 | α = .80 |
| Parent PROMIS PSRI | α = .88 | α = .90 | α = .89 |

**Note:** PROMIS PSD = PROMIS Pediatric Sleep Disturbances Short-Form; PROMIS PSRI = Patient-Reported Outcomes Measurement Information System Pediatric Sleep-related Impairment Short-Form; PROMIS EDDP = Patient-Reported Outcomes Measurement Information System Emotional Distress—Depression—Pediatric Scale; BPFSC-11 = Borderline Personality Features Scale for Children-11; DERS-SF = the Difficulties in Emotion Regulation Short-Form total scores; UPPS-P = the Urgency, Premeditation (lack of), Perseverance (lack of), Sensation Seeking, Positive Urgency, Impulsive Behavior Scale; SCIM = the Self-Concept and Identity Measure total scores; SIQ Jr. = the Suicidal Ideation Questionnaire Junior; ASHS = Adolescent Sleep Hygiene Scale; ASWS = Adolescent Sleep Wake Scale; CBCL = The Child Behavior Checklist.

**Additional Scoring information for Study Measures**

The *McLean Screening Instrument for Borderline Personality Disorder* (MSI-BPD; Zanarini et al., 2003), was initially developed as a 10-item yes/no self-report screening instrument to assess BPD features among individuals from diverse populations and has strong psychometric properties (i.e., Cronbach’s α = 0.74 for internal consistency; Spearman’s rho = 0.72 for trest-retest reliability; Zanarini et al., 2003). Our interview version employed stem questions from the original MSI-BPD, yet allowed study personnel to probe for examples and capture a more nuanced understanding of participants’ self-reported BPD features to ensure eligibility.

The *Patient-Reported Outcomes Measurement Information System Pediatric Sleep Disturbances* (PROMIS-PSD) short-form is an 8-item Likert-style self-report measure used to examine youth difficulties falling and staying asleep over the prior 7 days. The PROMIS *Pediatric Sleep-related Impairment* (PROMIS-PSRI) short-form uses 8 items to assess for daytime sleepiness and related impairment in the past week (e.g., difficulties concentrating, having fun, impaired productivity and poor mood due to sleepiness, troubles staying awake, etc.; Forrest et al., 2018). Each was validated in a large national sample (Forrest et al., 2018). Response options range from 1 (*never*) to 5 (*always*). Raw scores are converted to standardized t-scores.

The *Level 2 PROMIS Emotional Distress—Depression—Pediatric* Item Bank (PROMIS-EDDP; APA, 2013) is a 14-item Likert-style questionnaire assesses depression severity in the prior week. Response options range from 1 (*never*) to 5 (*always*). T-scores < 55 indicate no-to-slight depression, 55 to 59.9 reflect mild depression, 60 to 60.9 reflect moderate depression, and scores of 70+ indicate severe depression.

The *Difficulties in Emotion Regulation Scale Short Form* (DERS-SF) is an 18-item measure that yields a total score and six subscale scores (Kaufman et al., 2016). Respondents indicate how often items apply to them, with responses ranging from 1 (almost never) to 5 (almost always). The DERS-SF performs as well as the full original instrument (correlations between the two range from .90-.98 and reflecting 81-96% shared variance; Kaufman et al., 2016).

The *Urgency, Premeditation (lack of), Perseverance (lack of), Sensation Seeking, Positive Urgency, Impulsive Behavior Scale* (UPPS-P) assesses five impulsivity facets (Lack of Planning, Sensation Seeking, Negative Urgency, Positive Urgency, and Lack of Perseverance). Response options range from 1 (*agree strongly*) to 4 (*disagree strongly*). All responses were coded such that higher scores reflect higher impulsivity.

The *Self-Concept and Identity Measure* (SCIM; Kaufman et al., 2015) consists of 27 items on a 7-point Likert-scale (1 = *strongly disagree* to 7 = *strongly agree*), yielding a total score, and subscale scores for: consolidated identity, disturbed identity, and lack of identity. Consolidated identity items are reverse scored so that total SCIM scores indicate global difficulties. Prior work indicates that internal consistency of the SCIM is excellent (Cronbach’s α = .89), as is test-retest reliability (α = .93, *r* = .88; Kaufman et al., 2015). Internal reliability for the total scale in a prior adolescent sample was acceptable at .69, and the disturbed, consolidated, and lack of identity subscales were good to excellent (α = .83, .85, and .93 respectively; Kaufman et al., 2016).

The *Suicide Ideation Questionnaire Junior* (SIQ-Jr.; Reynolds, 1988) is a widely used Patient-Reported Outcome Measure designed to capture suicidal ideation in adolescent samples (Courtney et al., 2024). This 15-item questionnaire is used to assess the suicidal ideation frequency in the past month. Response options range from 0 (*I never had this thought*) to 6 (*almost every day*) and items are summed to yield a total score. A recent systematic review reported that the SIQ-Jr. demonstrates adequate to very good structural validity, and high internal consistency (Courtney et al., 2024).

The *Adolescent Sleep Hygiene Scale* (ASHS; LeBourgeois et al., 2005a) has response options that range from 1 (*Never, or 0% of the time*) to 6 (*Always, or 100% of the time*); whereas *the Adolescent Sleep Wake Scale* (ASWS; LeBourgeois et al., 2005b) items are rated on a 1 (*never*) to 6 (*always*) scale.

**Supplemental Ecological Momentary Assessment (EMA) Tables**

Table 1. *Wake Clock Time (Minutes from Midnight)*

|  | **Estimate** | **SE** | **95% CI** | **p-value** |
| --- | --- | --- | --- | --- |
| **Within Person Effects** |  |  |  |  |
| Wake Time ON Home | 84.27 | 8.70 | 67.23, 101.30 | < .001 |
| **Between Person Effects** |  |  |  |  |
| Slope WITH Wake Time | -1794.87 | 821.73 | -3405.47, -184.28 | .03 |
| Wake Time Mean | 455.59 | 10.74 | 434.54, 476.64 | < .001 |
| Slope Mean | -24.94 | 8.18 | -40.98, -8.91 | .002 |
| Wake Time Variance | 5091.29 | 1091.28 | 2952.37, 7230.21 | < .001 |
| Slope Variance | 2112.26 | 704.38 | 731.68, 3492.84 | .003 |

Table 2. *Sleep Clock Time (Minutes from Midnight)*

|  | **Estimate** | **SE** | **95% CI** | **p-value** |
| --- | --- | --- | --- | --- |
| **Within Person Effects** |  |  |  |  |
| Sleep Time ON Home | 37.20 | 9.12 | 19.33, 55.08 | < .001 |
| **Between Person Effects** |  |  |  |  |
| Slope WITH Sleep Time | -3310.74 | 904.37 | -5083.29, -1538.18 | < .001 |
| Sleep Time Mean | -16.88 | 11.73 | -39.87, 6.11 | .15 |
| Slope Mean | -47.78 | 11.39 | -70.10, -25.47 | < .001 |
| Sleep Time Variance | 6710.16 | 1224.66 | 4309.83, 9110.49 | < .001 |
| Slope Variance | 4254.50 | 1746.25 | 831.84, 7677.15 | .015 |

Table 3. *Time to Fall Asleep (Minutes)*

|  | **Estimate** | **SE** | **95% CI** | **p-value** |
| --- | --- | --- | --- | --- |
| **Within Person Effects** |  |  |  |  |
| MINUTES ON Home | -0.45 | 2.50 | -5.36, 4.46 | .86 |
| **Between Person Effects** |  |  |  |  |
| Slope WITH MINUTES | -213.60 | 231.12 | -666.60, 239.39 | .36 |
| MINUTES Mean | 33.66 | 3.75 | 26.32, 41.00 | < .001 |
| Slope Mean | -4.88 | 3.59 | -11.91, 2.16 | .17 |
| MINUTES Variance | 786.74 | 275.64 | 246.49, 1326.99 | .004 |
| Slope Variance | 249.10 | 183.78 | -111.10, 609.30 | .18 |

Table 4. *Number of Awakenings After Sleep Onset*

|  | **Estimate** | **SE** | **95% CI** | **p-value** |
| --- | --- | --- | --- | --- |
| **Within Person Effects** |  |  |  |  |
| Wakes ON Home | 0.04 | 0.12 | -0.20, 0.27 | .76 |
| **Between Person Effects** |  |  |  |  |
| Slope WITH Wakes | -0.13 | 0.13 | -0.38, 0.12 | .30 |
| Wakes Mean | 1.27 | 0.11 | 1.05, 1.49 | < .001 |
| Slope Mean | -0.25 | 0.11 | -0.47, -0.03 | 0.02 |
| Wakes Variance | 0.76 | 0.36 | 0.05, 1.47 | .04 |
| Slope Variance | 0.22 | 0.06 | 0.09, 0.34 | .001 |

Table 5. *Total Minutes Awake after Sleep Onset (Minutes)*

|  | **Estimate** | **SE** | **95% CI** | **p-value** |
| --- | --- | --- | --- | --- |
| **Within Person Effects** |  |  |  |  |
| MINUTES ON Home | 6.54 | 4.04 | -1.39, 14.46 | .11 |
| **Between Person Effects** |  |  |  |  |
| Slope WITH MINUTES | -47.92 | 120.76 | -284.62, 188.77 | .69 |
| MINUTES Mean | 23.49 | 3.63 | 16.37, 30.61 | < .001 |
| Slope Mean | -4.05 | 4.07 | -12.02, 3.92 | .32 |
| MINUTES Variance | 463.79 | 153.33 | 163.27, 764.32 | .002 |
| Slope Variance | 6.76 | 73.23 | -136.77, 150.28 | .93 |

Table 6. *Perceived Sleep Quality (1-5 Likert-Type)*

|  | **Estimate** | **SE** | **95% CI** | **p-value** |
| --- | --- | --- | --- | --- |
| **Within Person Effects** |  |  |  |  |
| Quality ON Home | -0.002 | 0.07 | -0.14, 0.14 | .98 |
| **Between Person Effects** |  |  |  |  |
| Slope WITH Quality | -0.19 | 0.08 | -0.35, -0.03 | .02 |
| Quality Mean | 3.15 | 0.09 | 2.97, 3.33 | < .001 |
| Slope Mean | 0.16 | 0.08 | -0.01, 0.32 | .06 |
| Quality Variance | 0.44 | 0.09 | 0.26, 0.62 | < .001 |
| Slope Variance | 0.21 | 0.09 | 0.04, 0.38 | .01 |

Table 7. *BPD Symptoms*

|  | **Estimate** | **SE** | **95% CI** | **p-value** |
| --- | --- | --- | --- | --- |
| **Within Person Effects** |  |  |  |  |
| BPD ON Home | -0.14 | 0.21 | -0.54, 0.27 | .51 |
| **Between Person Effects** |  |  |  |  |
| Slope WITH BPD | 0.28 | 0.23 | -0.18, 0.73 | .23 |
| Slope Mean | -1.57 | 0.34 | -2.23, -0.90 | < .001 |
| BPD Variance | 0.46 | 0.50 | -0.52, 1.44 | .36 |
| Slope Variance | 4.63 | 1.63 | 1.43, 7.84 | .005 |

Table 8. *Total Sleep Time (Minutes)*

|  | **Estimate** | **SE** | **95% CI** | **P Value** |
| --- | --- | --- | --- | --- |
| **Within Person Effects** |  |  |  |  |
| Total ON Home | 38.18 | 9.38 | 19.80, 56.57 | < .001 |
| **Between Person Effects** |  |  |  |  |
| Slope WITH Total | -2718.82 | 1020.13 | -4718.27, -719.36 | .008 |
| Total Mean | 434.19 | 11.08 | 412.47, 455.92 | < .001 |
| Slope Mean | 35.64 | 11.11 | 13.86, 57.41 | .001 |
| Total Variance | 5116.54 | 963.68 | 3227.73, 7005.35 | < .001 |
| Slope Variance | 3472.97 | 1330.73 | 864.74, 6081.21 | .009 |

Table 9. *Wish to live, wish to die*

|  | **Estimate** | **SE** | **95% CI** | **p-value** |
| --- | --- | --- | --- | --- |
| **Within Person Effects** |  |  |  |  |
| Wish to Live ON Home | 0.13 | 0.53 | -0.91, 1.17 | .81 |
| Wish to Die ON Home | -0.43 | 0.51 | -1.42, 0.57 | .40 |
| Wish to Live WITH Wish to Die | -169.28 | 32.11 | -232.22, -106.33 | < .001 |
| **Between Person Effects** |  |  |  |  |
| Live Slope WITH Wish to Live | -25.50 | 42.48 | -108.76, 57.76 | .55 |
| Live Slope WITH Wish to Die | 35.55 | 35.74 | -34.51, 105.61 | .32 |
| Live Slope WITH Die Slope | -87.62 | 34.05 | -154.36, -20.88 | .01 |
| Die Slope WITH Wish to Live | 17.73 | 41.43 | -63.47, 98.92 | .67 |
| Die Slope WITH Wish to Die | -7.09 | 36.54 | -78.70, 64.52 | .85 |
| Wish to Live WITH Wish to Die | -425.39 | 73.10 | -568.67, -282.10 | < .001 |
| Wish to Live Mean | 73.73 | 2.90 | 68.05, 79.40 | < .001 |
| Wish to Die Mean | 19.69 | 2.49 | 14.81, 24.58 | < .001 |
| Live Slope Mean | 1.93 | 1.72 | -1.43, 5.30 | .26 |
| Die Slope Mean | -1.03 | 1.51 | -3.99, 1.92 | .49 |
| Wish to Live Variance | 578.69 | 80.20 | 421.50, 735.89 | < .001 |
| Wish to Die Variance | 410.34 | 73.06 | 267.15, 553.53 | < .001 |
| Live Slope Variance | 143.04 | 41.84 | 61.02, 225.05 | .001 |
| Die Slope Variance | 104.56 | 35.14 | 35.69, 173.44 | .003 |

Table 10. *Negative Affect (NA) and Positive Affect (PA).*

|  | **Estimate** | **SE** | **95% CI** | **p-value** |
| --- | --- | --- | --- | --- |
| **Within Person Effects** |  |  |  |  |
| NA ON Home | -0.73 | 0.64 | -1.98, 0.52 | .25 |
| PA ON Home | 1.42 | 0.83 | -0.21, 3.06 | .09 |
| NA WITH PA | -122.37 | 13.92 | -149.65, -95.09 | < .001 |
| **Between Person Effects** |  |  |  |  |
| NA Slope WITH NA | -43.43 | 26.50 | -95.38, 8.51 | .10 |
| NA Slope WITH PA | 39.47 | 26.03 | -11.54, 90.48 | .13 |
| NA Slope WITH PA Slope | -48.77 | 25.38 | -98.52, 0.98 | .06 |
| PA Slope WITH NA | 48.32 | 22.72 | 3.78, 92.85 | .03 |
| PA Slope WITH PA | -32.63 | 23.05 | -77.81, 12.55 | .16 |
| NA WITH PA | -103.22 | 37.33 | -176.38, -30.06 | .006 |
| NA Mean | 29.24 | 2.15 | 25.02, 33.46 | < .001 |
| PA Mean | 50.15 | 2.08 | 46.08, 54.22 | < .001 |
| NA Slope Mean | -3.95 | 1.84 | -7.56, -0.35 | .03 |
| PA Slope Mean | 0.91 | 1.53 | -2.09, 3.90 | .55 |
| NA Variance | 302.17 | 44.72 | 214.53, 389.81 | < .001 |
| PA Variance | 285.96 | 50.72 | 186.54, 385.38 | < .001 |
| NA Slope Variance | 167.70 | 49.25 | 71.17, 264.22 | .001 |
| PA Slope Variance | 112.81 | 22.56 | 68.60, 157.02 | < .001 |

Table 11. *Arguments*

|  | **Estimate** | **SE** | **95% CI** | **P Value** |
| --- | --- | --- | --- | --- |
| **Within Person Effects** |  |  |  |  |
| Argue ON Home | 0.13 | 0.17 | -0.20, 0.46 | 0.43 |
| **Between Person Effects** |  |  |  |  |
| Slope WITH Argue | 0.10 | 0.38 | -0.65, 0.85 | 0.80 |
| Argue Threshold | 2.85 | 0.22 | 2.43, 3.28 | < .001 |
| Slope Mean | -0.80 | 0.27 | -1.33, -0.27 | .003 |
| Argue Variance | 1.48 | 0.36 | 0.77, 2.18 | < .001 |
| Slope Variance | 0.01 | 0.07 | -0.13, 0.15 | 0.90 |

**Participant enrollment by month**

| **Month** | **Participants Enrolled** |
| --- | --- |
| January | 7 |
| February | 5 |
| March | 8 |
| April | 0 |
| May | 1 |
| June | 6 |
| July | 4 |
| August | 5 |
| September | 1 |
| October | 7 |
| November | 8 |
| December | 5 |
